# Association between epidemic dynamics of Covid-19 infection and ABO blood group types

**DOI:** 10.1101/2020.07.12.20152074

**Authors:** Yuefei Liu, Lisa Häussinger, Jürgen M. Steinacker, Alexander Dinse-Lambracht

**Author notes:** For correspondence: Prof. Dr. Yuefei Liu, Division of Sports and Rehabilitation Medicine, Department of Internal Medicine II, University of Ulm, Ulm. Germany, Leimgrubenweg 14, D-89075 Ulm. Germany.

## Abstract

**Background:** Covid-19 pandemic is the most critical challenge nowadays for the manhood, and the infection and death cases are still speedily increasing. Since there are no available vaccine and specifically effective treatment, to break the infectious way of the pandemic remains the unique measure to efficiently combat Covid-19 infection. Understanding factors that affect the Covid-19 infection can help make better balance between activity restriction and infection dynamics. This study sought to investigate association between Covid-19 infection and blood type distribution.

**Methods:** The big data provided by World Health Organization and Johns Hopkins University were taken to assess epidemic dynamics of Covid-19 infection. Growth rate and doubling time of infection and death cases, reproductive number, infection and death cases in the mid-exponential phase were analyzed in relation to blood type distribution.

**Results:** Growth rate of infection and death cases correlated significantly to blood type A proportion of the population positively while to blood type B proportion negatively. In comparison with lower blood type A population (< 30%) people with higher blood type A (≥ 30%) had more infection and death cases in the early exponential phase, higher growth rates, and shorter case doubling time for infection and death.

**Discussion:** Covid-19 infection is significantly associated with blood type distribution and people with blood type A are more susceptible to Covid-19 infection and have higher epidemic dynamics and higher case fatality rate. The results of this study provide important and useful information for fighting Covid-19 pandemic.

## Introduction

The pandemic of Covid-19 infection still keeps rapidly increasing worldwide, which threatens very much the public health and causes dramatic knockdown of the global economics and social life. According to the data derived from World Health Organization (WHO) database, up to date there was over 10 million confirmed cases of Covid-19 infection (WHO situation reports on June 29, 2020), and the daily new infection cases maintain at very high level. To control this pandemic outbreak is difficult because factors affecting the Covid-19 infection have yet not been thoroughly understood, and specific vaccine and treatments are still unavailable.

If one looks the Covid-19 infection map provided by Johns Hopkins University (JHU) (for example the map picture on May15, 2020), one would immediately recognize that there is quite a difference among the geographic districts. In fact, after the initial outbreak in China, the Covid-19 pandemic speedily advanced to Europe and simultaneously to New York City, and afterwards this pandemic spread to South America and East Mediterranean zones. Trying to understand the factors that have profound impact on the pandemic is crucial for bringing the pandemic under control since factors associated with the pandemic must be considered to make public policy and medical decisions. ABO blood types are attributed to diverse infectious diseases like malaria, HIV and influenza (1-4). It has been reported that among the confirmed Covid-19 infection cases that were treated in the hospitals the proportion of blood type A was significantly higher than that of blood type B (5), and furthermore it has been reported that the severity and clinical outcome of the Covid-19 infection disease were associated with blood types (6).

However, in these previous studies the study subjects were in a relatively small number and/or limited locality so that the data cover only a regional geographic zone and do not reveal the worldwide geographically uneven distribution of Covid-19 infection. Furthermore, the dynamics of Covid-19 had not been dealt with worldwide. We therefore conducted this big-data-analysis on association between dynamics of Covid-19 pandemic and ABO blood types. The big data are derived from the official database presented by World Health Organization (WHO). ABO blood type distribution serves as a typical genetic marker for geographic distribution over the globe for diverse diseases as well as public health issues. Finding out any factors that are attributed to Covid-19 infection might be thus important in fighting Covid-19 pandemics.

For an epidemic of an infectious disease the dynamic development of the infection is determinant, and this can be assessed by several parameters classically used, that are among others infection case growth rate (ICGR), infection case doubling time (IC-dt) reproductive number (RN), death case growth rate (DCGR), and death case doubling time (DC-dt). The difficulty to determine these parameters is that the current high dynamic in the infection development worldwide so that an endpoint of total infection number remains yet unreached. Instead, we tried to assess on the mid-way of the exponential phase of infection the infection cases and death cases, which was performed based on the epidemical curves of the involved countries displayed by JHU.

This study sought to investigate the relationship between the distribution of blood group types and the epidemic dynamics of Covid-19 infection based on analyses of big data that cover worldwide population majority.

## Methods

For analysis of the pandemic of Covid-19 infection the population of six geographic regions divided according to WHO is included: Africa, America, East Mediterranean, Europe, South East Asia and Western pacific. Within in each geographic zone six countries are randomized selected for the analysis (table 1 supplement material). However, at the time point of the analysis two countries, i.e. Botswana and Papua New Guinea, got only a few Covid-19 infection cases that did not show an epidemic and therefore were excluded from the analysis. Finally, the total population of the 34 countries was about 5391149 thousand (2016 data of WHO).

Data of blood type distribution of each country are derived from original research studies with respect to each corresponding countries (table 1 of supplement material). Since the blood type A seemed more relevant through prior correlation analysis (figure 2a), and mean blood type A proportion of all 34 countries was 30%, the population of all included countries was divided in to higher (≥ 30%) and lower (<30%) blood type A groups for advanced analysis (figure 3).

The confirmed infection cases as well as death cases of each country were taken from the daily situation reports of WHO. To calculate ICGR the start point (1^st^ day) was set on the day when the infection cases began to increase exponentially. This point could be identified on the curve of JHU by two independent coauthors. Taking Germany as an example, the day when the infection cases began to increase was March 12, 2002 with 1567 infection cases. The afterward-consecutive 14 daily cases were input into SPSS^®^. ICGR was calculated according to the following formula in the SPSS software:

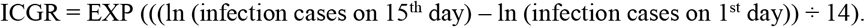

The calculation of RN was referred to the methods used by Robert-Koch-Institute (Cologne. Germany) (6-8). The calculation was based on the assumption of the viral generation time with 4 days and the incubation time 5 days. According to the following formula, the RN was calculated:

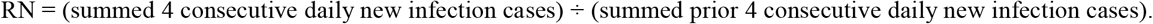

The death cases caused by or with Covid-19 infection were taken from the daily situation report of WHO. Based on the clinical consideration that a death case took place about 15 days after infection, the death cases on the 16^th^ day and the following consecutive 14 days were input into SPSS^®^ to calculate the death case growth rate (DCGR) that was calculated analogously to that of ICGR as following:

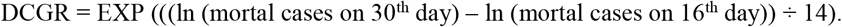

Furthermore, the doubling time was calculated for infection cases (IC-dt) and for death cases (DC-dt), respectively as follows:

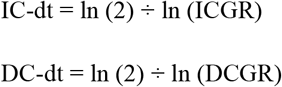

From the epidemical curve provided by JHU, a plateau phase for the infection cases could be considered when the increase of new infection cases was no longer exponential, which could be assessed for a part of the countries. The time interval from the start point of exponential phase to the begin of the plateau phase could be determined (days to begin of the plateau phase, Dpp). Furthermore, the cases on the day when the plateau phase was reached (nine countries up to June 30, 2020) were taken for calculation of Covid-19 infection of the corresponding country. The mean Dpp was 51.2 days among these nine countries and thus Dpp_(1/2)_ was set at 26 days after begin of the exponential phase. Since most of the analyzed countries did not reach their plateau phase, Dpp_(1/2)_ was taken for all analyzed countries to count the infection and death cases (IC_Dpp(1/2)_ and DC _Dss(1/2)_, respectively). All these methodological measures are depicted in Figure 1.

**Figure 1.**
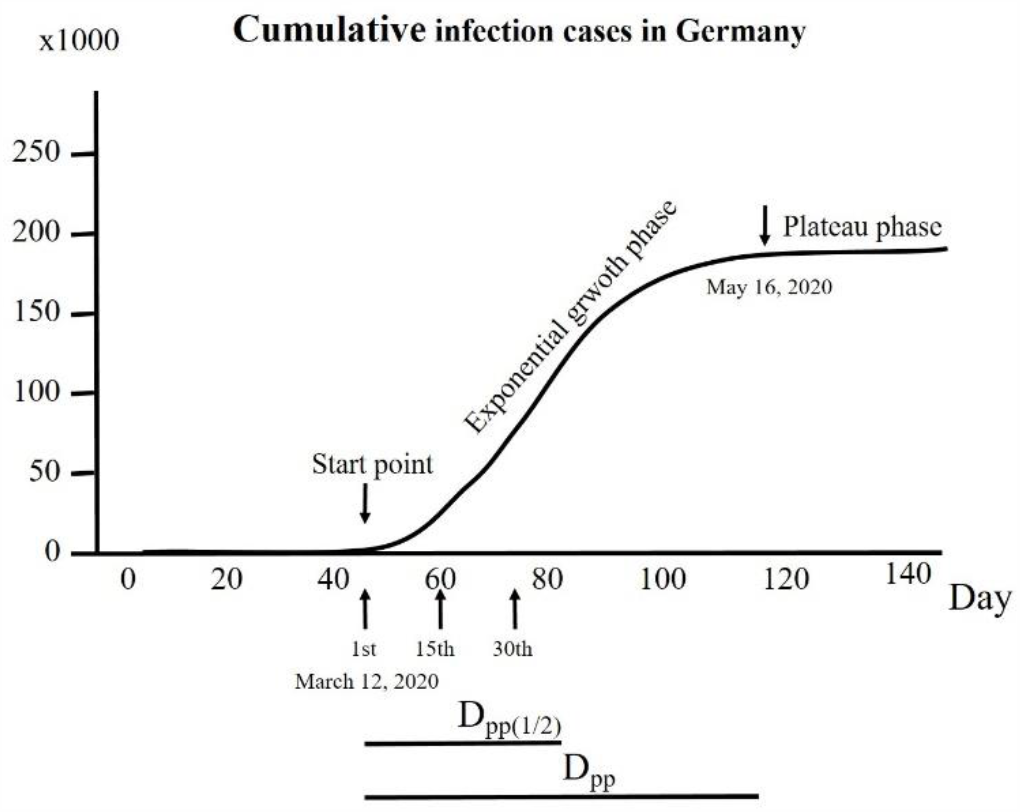
Illustration of mathematical analyses of the study parameters involving Covid-19 infection. As an example the development of infection cases in Germany over 140 days is displayed to determine the points taken for calculations. From the course of cumulative infection cases a curve with different phases can be identified. The 1^st^ day was set on the day when the infection cases began to increase exponentially. In the early exponential phase the infection growth rate (ICGR) was calculated. The death cases were taken from the period day14 days after the start point through further 14 days. The plateau phase was reached when the course of cumulative infection cases did no longer show an exponential increase. The time interval (days) between start point and begin of plateau phase (Dpp) was calculated. For in most of the countries a plateau phase was yet not reached, Dpp_(1/2)_ was set as half mean Dpp and calculated for all the countries.

In addition, a comparison of the epidemic dynamics was performed by dividing the population into higher and lower groups of blood type A proportion either with nonparametric (Mann Whitney) (figure 3a) or ANOVA (figure 3b) accordingly. Because the mean proportion of blood type A was 30%, the higher and lower blood type A groups were defined respectively by ≥ 30% or < 30%.

The mathematic and statistical procedures were performed with SPSS^®^ (IBM. 25^th^ edition. USA). A difference was assumed to be significant by P<0.05.

## Results

In table 2 are summarized the parameters describing the Covid-19 epidemic dynamics, an overall difference among the selected countries was found for each parameter through ANOVA analyses (P<0.01).

ICGR correlates positively with blood type A (R^2^ = 0.328, P<0.05. Figure 2a) and negatively with type B (R^2^ = 0.210, P<0.05. Figure 2b), but not with blood type O or AB.

**Figure 2.**
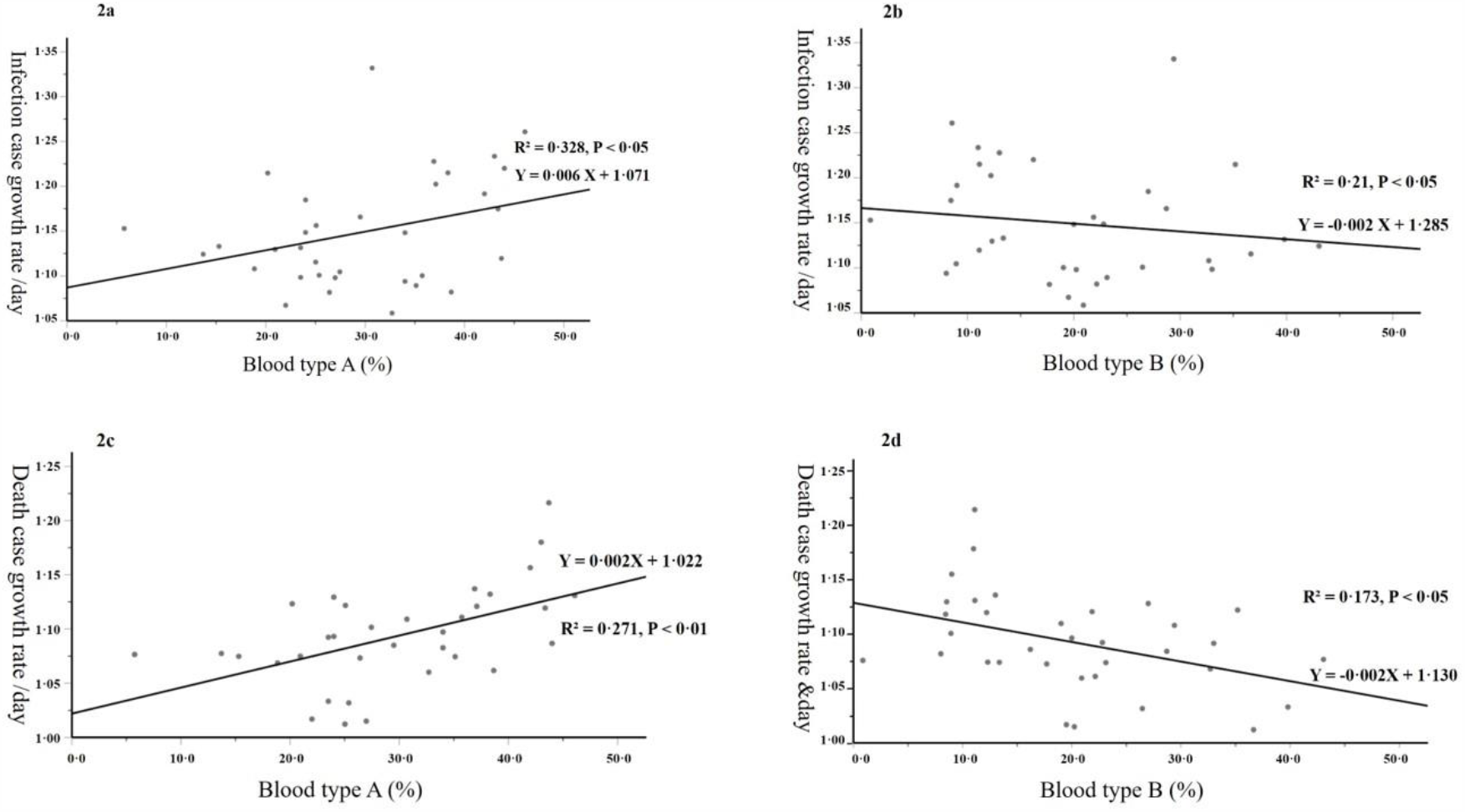
Correlation between blood type distribution and Covid-19 infection case growth rate per day and death case growth rate per day.

The relationship between DCGR and blood types is shown in Fig.3. DCGR correlated positively with blood type A (P < 0.01, Figure 2c) and negatively with blood type B (P < 0.05, Figure 2d), but not with blood type O or AB.

**Figure 3.**
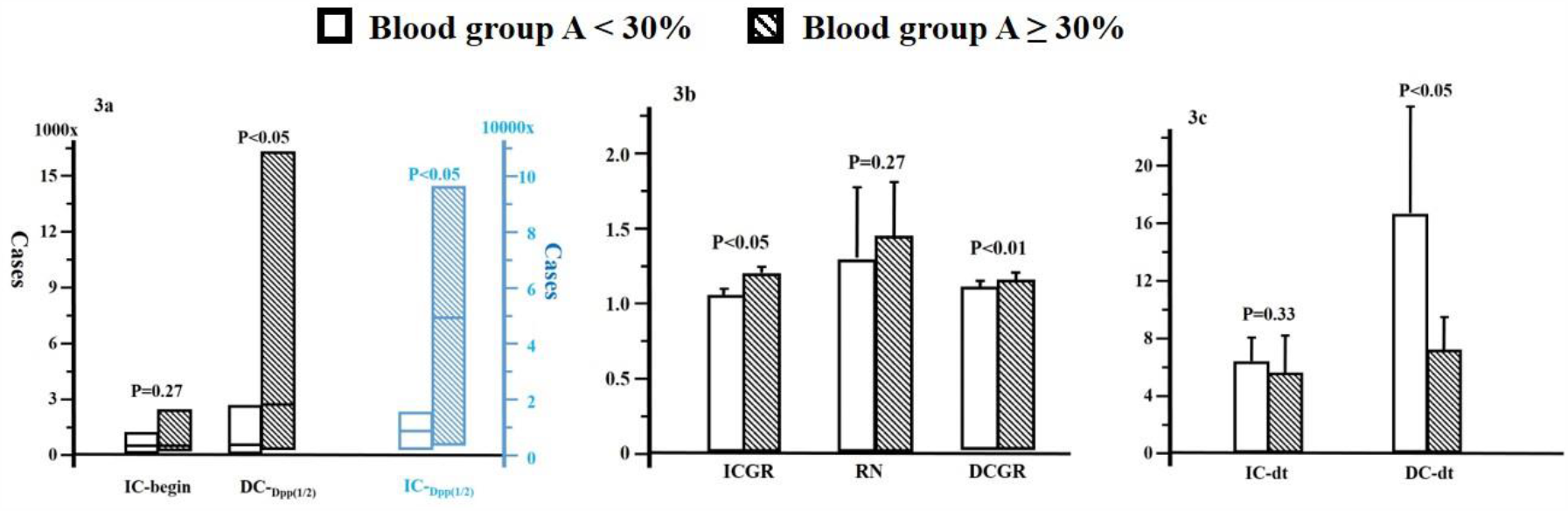
Comparison of the Covid-19 infection epidemic dynamics between higher and lower blood type A population. In figure 3a: IC-begin, the first day when infection case began to increase exponentially; DC_Dpp(1/2)_, death cases on 26^th^ day after IC-begin; IC_Dpp(1/2)_, infection cases on 26^th^ day after IC-begin. In figure 3b: ICGR, infection case growth rate per day; RN, reproductive number; DCGR, death case growth rate per day. In figure 3c: IC-dt, infection case doubling time in day; DC-dt, death case doubling time in day.

At begin of exponential phase of Covid-19 infection, the infection cases were comparable between the higher and lower blood type A groups (P > 0.05, figure 3a), but at Dpp_(1/2)_ the infection cases were distinctly higher in group with higher blood type A than that with lower blood type A (P < 0.05), which was also the case for to DC_Dpp(1/2)_ (P < 0.05).

During the first 15 days of exponential growth phase ICGR was significantly higher in the group with higher blood type A than that in the group with lower blood type A (P < 0.05, Figure. 3b). IC-dt was reciprocally shorter in the higher blood group A than that of the lower blood type group A (P < 0.05, figure 3c). Though the reproductive number showed slightly higher in the group with higher blood type A than that in the lower blood group A, the difference was statistically not significant (P > 0.05, Figure 3b).

The dynamic changes in death cases indicated by DCGR showed significant difference between both groups, i.e. higher in the group with higher blood type A proportion (P < 0.01, Figure 3b). Accordingly, the death case doubling time was significantly shorter in this group (P < 0.05, Figure 3c).

## Discussion

The worldwide pandemic of Covid-19 infection is dramatically challenging the global health care, social life and economy, and is spreading with still speedy increase in new infection and death cases. To control and conquer this pandemic is of critical interest for the manhood and global society.

Classically, to control an epidemic disease, four major measures are essential, they are controlling disease origin, cutting the infectious way, effectively treating patients and improving herd immunity. However, up to date, the origin of Covid-19 epidemic remains uncertain; a clinically proven vaccine to improve immunity against Covid-19 virus is still unavailable; an effective therapy for Covid-19 infected patients has yet not established. Therefore, the unique certain effective measure at present is to break the way of wide spreading, i.e., activity restriction and community lockdown. Unfortunately, lockdown causes a series of adverse effects, e.g., down of community economy, reduction of productivity, restriction of people freedom, and hamper of the public health care. Therefore, to balance rationally the epidemic dynamics and lockdown is of critical social and health interest, which bases on the observation and analysis of Covid-19 infection dynamics.

To determine finally the dynamics of Covid-19 infection is by no means easy because a number of factors have profound impacts on this dynamics. It is hardly possible to get the exact data of infection and death cases due to limited manpower and health care resource in testing the population, not to mention limited test certainty in terms of sensitivity as well as specificity, variant incubation time, different social environments and activities, etc. Therefore, analysis based on big data seems a reasonable way to assess the dynamics of Covid-19 infection globally. At present, WHO and JHU are monitoring this dynamics and providing useful information, we therefore take these useful tools and go further with analysis with specific interest on genetic factor.

In fact, genetic factor plays important role in many diseases like cardiovascular diseases (9), diabetes mellitus (10), cancers (11),, and infectious diseases including malaria (12), HIV and influenza (13;14). As a genetic factor, ABO blood type distribution has been extensively investigated in the field of infectious diseases (13-15). Meanwhile, investigation interest on relationship between ABO blood system and Covid-19 infection is increasingly (6;16;17). The first study suggested that among the infected individuals the proportion of blood type A patients was higher (17). In this study, total 2173 confirmed Covid-19 infection cases treated in hospitals were analyzed, and of these patients a higher population with blood group A than that of comparable population without Covid-19 infection was found. Since the study subjects were in a small number and from the same locality, the data were quite limited. A larger multicenter study has been recently reported by Ellinghaus et al, and a distinct association between ABO blood types and severity of Covid-19 infection disease could be demonstrated (6;16;17). Unfortunately, in their study the dynamics of the Covid-19 infection was not investigated. We thus conducted this study based on the worldwide available big data trying to ascertain an association between the distribution of blood types and Covid-19 infection dynamics.

The results of our analyses show that there was a positive correlation between proportion of blood group A in the worldwide population and the dynamics of Covid-19 infection reflected by the infection case growth rate, and a negative correlation was found between blood group B and the infection dynamics (Figure 2). Furthermore, similar results could be also elucidated for the relationship between blood group types and death case growth rate (Figure 2). In the context, our results are in a good accordance to these aforementioned studies, and strongly demonstrate the association between the distribution of blood group types and Covid-19 infection.

Based on the above described results we divided the worldwide population into two groups according to the proportion of blood type A, i.e., higher blood type A group (≥30%) and lower group (<30%), and compared the differences in terms of the epidemic dynamic parameters between both the groups. It is shown in Figure 3 that in the higher blood type A group except for the infection cases at begin of exponential phase and the reproductive number, all other parameters were significantly higher than those in the lower blood type A group. These results demonstrate further that an association between blood type distribution and the epidemic dynamics of Covid-19 infection. We have also analyzed a relationship between the population life expectance as well as health care expense and Covid-19 infection cases (data not shown) based on data provided by WHO. That with increase in life expectance the Covid-19 infection cases per 100,000 people increased, suggesting that the older people have a higher Covid-19 infection susceptibility, is already well-known. There was no significant correlation between health care expense and Covid-19 infection cases in the analyzed countries.

All these results mentioned above strongly demonstrate that the epidemic dynamics of Covir-19 infection is associated with the distribution of blood types worldwide, not only infection cases but also death cases. People with blood group A are therefore more susceptible to Covid-19 infection and their case fatality rate though Covid-19 infection is higher.

Mechanisms responsible for this association are yet to be explored. Nowadays, it is believed that Covid-19 infection begins with the docking of its spiking protein to ACER2 (18-20). Whether ACER2 expression level is different among the blood types, and whether people with blood type A have higher expression level of ACER2, remain unclear. Certainly, further studies are necessary to explore the mechanisms.

Bearing the association between Covid-19 infection and blood type distribution in mind may important and meaningful in order to make reasonable decision in combating the Covid-19 infection among different people with their blood type distribution. In general, countries with lower proportion of blood type A have lower economic income and poorer health care resource, which might be exaggerated by community lockdown. It might reasonable for these countries to make slower or later strict measures owing to the lower epidemic dynamics of Covid-19 infection, whereas for those countries where the people with higher proportion of blood type A the strict measure to combat the Covid-19 infection ought to be more active and earlier.

## Data Availability

World Health Organization
Johns Hopkins University

https://www.who.int

https://coronavirus.jhu.edu/map.html

## References

(1) Damena D, Denis A, Golassa L, Chimusa ER. Genome-wide association studies of severe P. falciparum malaria susceptibility: progress, pitfalls and prospects. BMC Med Genomics 2019;14:120. doi: 10.1186/s12920-019-0564-x.

(2) Degarege A, Gebrezgi MT, Ibanez G, Wahlgren M, Madhivanan P. Effect of the ABO blood group on susceptibility to severe malaria: A systematic review and meta-analysis. Blood Rev 2019;33:53–62.

(3) Siransy LK, Nanga ZY, Zaba FS, Tufa NY, Dasse SR. ABO/Rh Blood Groups and Risk of HIV Infection and Hepatitis B Among Blood Donors of Abidjan, Cote D’ivoire. Eur J Microbiol Immunol 2015;5:205–9.

(4) Horby P, Nguyen NY, Dunstan SJ, Baillie JK. The role of host genetics in susceptibility to influenza: a systematic review. PLoS One 2012;7:e33180.

(5) Wu Y, Feng Z, Li P, Yu Q. Relationship between ABO blood group distribution and clinical characteristics in patients with COVID-19. Clin Chim Acta 2020;509:220–3.

(6) Ellinghaus D, Degenhardt F, Bujanda L, Buti M, Albillos A, Invernizzi P et al. Genomewide Association Study of Severe Covid-19 with Respiratory Failure. N Engl J Med 2020; DOI: 10.1056/NEJMoa2020283.

(7) an der Heiden M, Hamouda O. Schätzung der aktuellen Entwicklung der SARS-Cor-2-Epidemie in Deutschland - Nowcasting. Epid Bull. 2020;17:10–6

(8) Zietz M, Tatonetti NP. Testing the association between blood type and COVID-19 infection, intubation, and death. medRxiv 2020; doi: https://doi.org/10.1101/2020.04.08.20058073.

(9) Capuzzo E, Bonfanti C, Frattini F, Montorsi P, Turdo R, Previdi MG et al. The relationship between ABO blood group and cardiovascular disease: results from the Cardiorisk program. Ann Transl Med 2016;4:189–94.

(10) Williams DR, Cartwrigth RA. Genetic polymorphisms in diabetics and non-diabetics. J Med Genet 1979;16:351–7.

(11) Antwi SO, Bamlet WR, Pedersen KS, Chaffee KG, Risch HA, Shivappa N et al. Pancreatic cancer risk is modulated by inflammatory potential of diet and ABO genotype: a consortia-based evaluation and replication study. Carcinogenesis 2018;39:1056–67.

(12) Alemu G, Mama M. Assessing ABO/Rh Blood Group Frequency and Association with Asymptomatic Malaria among Blood Donors Attending Arba Minch Blood Bank, South Ethiopia. Malar Res Treat 2016; http://dx.doi.org/10.1155/2016/804376.

(13) Tyrrell DA, Sparrow P, Beare AS. Relation between blood groups and resistance to infection with influenza and spome picornaviruses. Nature 1968;220:819–20.

(14) Davison GM, Hendrickse HL, Matsha TE. Do Blood Group Antigens and the Red Cell Membrane Influence Human Immunodeficiency Virus Infection? Cells 2020;9:845–55.

(15) Evans AS, Shepard DA, Richards VA. ABO blood groups and viral diseases. Yale J Biol Med 1972;45:81–92.

(16) Zaidi FZ, Zaidi ARZ, Abdullah SM, Zaidi SZA. COVID-19 and the ABO blood group connection. Transfus Apher Sci 2020; ID: covidwho-505545.

(17) Zhao J, Yang Y, Huang H, Li D, Gu D, Lu X et al. Relationship between the ABO blood groups and the Covid-19 susceptibility. medRxiv 2020; doi: https://doi.org/10.1101/2020.03.11.20031096.

(18) Guillon P, Clemnet M, Sebille V, Rivain JG, Chou CF, Ruvoen-Clouet N et al. Inhibition of the interaction between ths SARS-CoV spike protein and its cellular receptor by anti-histo-blood group antibodies. Glyconbiology 2003;18: 1085–93.

(19) Wan Y, Shang J, Graham R, Baric RS, Li F. Receptor recognition by novel coronavirus from Wuhan: An analysis based on decade-long structural studies of SARS. J Virol 2020;94:DOI: 10.1128/JVI.00127-20.

(20) Hoffmann M, Kleine-Weber H, Schroeder S, Krüger N, Herrler T, Erichsen S et al. SARS-CoV-2 cell entry depends on ACE2 and TMPRSS2 and is blocked by a clinically proven protease inhibitor. Cell 2020;181:271–80.

